# (Mis) Communicating The Gut Microbiome: A Cross-Sectional Assessment of Social Media Video Content

**DOI:** 10.1101/2022.01.16.22269387

**Authors:** S Chidambaram, Y Maheswaran, C Chan, L Hanna, H Ashrafian, SR Markar, V Sounderajah, J Alverdy, A Darzi

**Affiliations:** Department of Surgery & Cancer, Imperial College London, London, United Kingdom; Institute of Global Health Innovation, Imperial College London, London, United Kingdom; Department of Surgery, University of Chicago, Illinois, USA

**Keywords:** microbiome, social media, YouTube, misinformation, content analysis

## Abstract

**Background:** Social media platforms, such as YouTube, are an integral means of disseminating information about health and wellness to the public. However, anecdotal reports have cited that the human gut microbiome has been a particular focus of dubious, misleading and, on occasion, harmful media content. Despite these claims, there have been no published studies investigating this phenomenon within popular social media platforms.

**Aim:** This study aims to (1) evaluate the accuracy and credibility of YouTube videos related to the human gut microbiome and (2) investigate the correlation between content engagement metrics and information quality, as defined by validated criteria.

**Methods:** In this cross-sectional study, videos about the human gut microbiome were searched on the UK version of YouTube on 20^th^ September 2021. The 600 most viewed videos were extracted and screened for relevance. Information content and characteristics were extracted and independently rated using the DISCERN quality criteria by two researchers.

**Results:** Overall, 319 videos accounting for 62,354,628 views were included. 73.4% of the videos were produced in North America, and over 78.6% were uploaded between 2019-2021. 41.0% of videos were produced by non-profit organizations. 16% of the videos had an advertisement for a product or promoted a health-related intervention for financial purposes. Videos by non-medical educational creators had the highest total and preferred viewership. Daily viewership was highest for videos by internet media sources. The average DISCERN and HONcode scores were 49.5 (0.68) out of 80 and 5.05 (2.52) out of 8, respectively. DISCERN scores for videos by medical professionals (53.2 (0.17)) were significantly higher than videos by independent content creators (39.1 (5.58)), (p<0.01). Videos with promotional material scored significantly lower DISCERN scores than videos without any advertisements or product promotion (p<0.01). There was no correlation between DISCERN scores and total viewership, daily viewership or preferred viewership (number of likes).

**Conclusions:** The overall credibility of information about human gut microbiome on YouTube is poor. Moreover, there was no correlation between the video information quality and the level of public engagement. The significant disconnect between reliable sources of information and the public suggests that there is an immediate need for cross-sector initiatives in order safeguard vulnerable viewers from the potentially harmful effects of misinformation and disinformation.

## 1. Introduction

Over the past decade, governments, health organisations and private sector companies have increasingly turned to the use of social media platforms as a trusted and official means of disseminating information on health and wellness (1–3). Of the available platforms, YouTube has arguably emerged as the standout modality for accessing comprehensive healthcare information. As of December 2021, YouTube boasts over one billion hours’ worth of watched content daily; over 2 billion unique users monthly; and is commonly accessed by both health professionals and the general public alike for a wide range of health related queries (4). Although YouTube’s usually hosts factually accurate content, there are instances in which inaccurate information has been hosted for public viewing (5). This issue came to prominence during the COVID-19 pandemic as inaccurate content about the vaccination programme rapidly gained global viewership. This, in turn, led to YouTube’s implementation of a COVID-19 medical misinformation policy, which disallowed COVID-19 content that contradicts the policies of health authorities (6). Since its implementation in September 2020, the platform has removed over 130,000 videos, a staggering figure highlighting the scale of misinformation on vital public health matters. However, a similarly specific policy has not been extended to other areas of health-related content in which there may be history of misinformed content. In these cases, the responsibility of determining the veracity and applicability of the encountered information is placed upon the individual (7).

As increasing understanding develops around the ‘digital determinants of health’ (8), there is concern that unverified content may lead to instances of clinical harm to viewers who cannot independently discern content quality (9–11). Moreover, these users may also share misinformed content to their own social networks, resulting in misinformation within epistemic bubbles and, more concerningly, echo chambers (12,13). One topic that is highly vulnerable to this phenomenon is the human gut microbiome. The gut microbiome is a consistently popular topic for content creators in YouTube, particularly as greater understanding develops around its regulatory role on anxiety, mood, cognition, and pain through the gut– brain axis. Despite the paucity of high-grade evidence on the mechanisms through which the microbiome may be reconfigured to optimise human physiology, there has been a sharp rise in videos that promote lifestyle-based strategies which purport to modify the gut microbiome for health and wellbeing benefits (14,15). Without rigorous standards related to the scientific accuracy, these videos often stretch the applications of existing scientific evidence to advocate treatments that are advertised as helpful but instead may have no effect or potentially cause overall harm. Hence, if not properly caveated with scientific resources and disclaimers, these videos can distract viewers away from empirically sourced medical advice and protective health seeking behaviours.

To date, there have been no studies which have evaluate the credibility of microbiome related content available on social media platforms. Moreover, it is not known whether the engagement metrics associated with microbiome related content correlates with the information credibility. As such, this study aims to (1) evaluate the accuracy and credibility of YouTube videos on the information provided about the human gut microbiome and (2) investigate the correlation between social media engagement metrics and information quality, as defined by validated criteria.

## 2. Methods

### 2.1. Selection of videos

The phrases “microbiome”, “gut microbiome” and “microbiome health” were searched on the UK version of YouTube on September 20, 2021. The search terms were chosen to ensure appropriate coverage of any videos related to the role of the gastrointestinal microbiome in health and well-being. The search was conducted in an incognito browser (Google Chrome) to avoid biased suggestions based on cookies or previous search history. The results were ranked according to view count as this is the most sensitive means of identifying videos that have had the greatest impact and are most likely to trend. The 200 most viewed videos (10 pages) of each search were de-duplicated and subsequently extracted. Video titles and channels were first screened for relevance and English language before further full-video screening. Videos were included if they described at least one of the following: (1) components of the gut microbiome; (2) role of microbiome in gut health including cancers, inflammatory bowel conditions, infectious bowel conditions; (3) methods of altering the gut microbiome for the purpose of broader health-related effects; and (4) side effects or safety of interventions used. Videos that did not discuss the human gut microbiome or were not in the English language were not included. In cases of uncertainty on whether a video should be included, a consensus was sought between authors, with a predisposition to include the video for full assessment.

### 2.2. Data extraction

Characteristics of the videos were independently extracted by two authors (SC and YM). This included the video’s Uniform Resource Locator (URL), channel name, country of origin, duration and age of video. Engagement metrics that assess the use of video by the public were also extracted; this included the number of likes, dislikes, comments and view count. Videos were subsequently classified based on their content and purpose into six categories: educational channels produced by medical professionals (e.g., the American Society for Microbiology); educational channels produced by nonmedical individuals (science education or explanatory media e.g., The Sheekey Science Show), independent producers (e.g., Tom Bilyeu – QUEST nutrition), internet media (magazines or talk shows e.g., Health Via Modern Nutrition), news agencies (clips from network news e.g., King 5 news), and non-profit or medical organizations (hospitals, government organizations, or universities e.g., Mayo Clinic, UCLA Health, Hopkins Kimmel).

### 2.3. Outcome measures

The primary outcome measure was the information quality of videos assessed, which is graded using the DISCERN tool, as well as adherence to the HONcode. Secondary measures included engagement metrics which were classified as total viewership (total number of views the videos accrued), daily viewership and preferred viewership (number of “likes” received by the video). Correlations between video characteristics, engagement metrics and quality assessment scores were analysed using linear regression models.

### 2.4. The DISCERN Score

The DISCERN tool is an instrument that is designed to help users assess consumer health information. The criteria were originally drafted based on the analysis of a random sample of consumer health information on the treatment choices for three medical conditions (myocardial infarction, endometriosis and chronic fatigue syndrome). The tool consists of three categories of items that assess the reliability (eight questions), quality of information (seven questions) and general impression (one question) of the information source. Each item is scored on a scale of 1 (worst) to 5 (best), accounting for a total score of 80. The DISCERN criteria has enabled the assessment of various aspects of information quality, including its completeness, understandability, relevance, depth, and accuracy of information. The shortlisted videos were rated independently by two authors (SC and YM). Any discrepancies were resolved through discussion with a third author (VS).

### 2.5. The Health on the Net Foundation Code of Conduct (HONcode)

The Health on the Net Foundation Code of Conduct (HONcode) certification is an ethical standard aimed at offering quality health information. It demonstrates the intent of a website to publish reliable and good-quality information with maximum transparency. The HONcode consists of eight principles that evaluate the reliability and credibility of health information, including justification and balance of claims; citation of sources used; details of funding; and clear distinguishment of advertising from editorial content (16). Videos were rated with a score of 1 (adherent) or 0 (nonadherent) for each of the 8 principles, for a total score of 8.

### 2.6. Statistical analysis

Statistical analysis was performed using SPSS (version 27, IBM Corporations). All data are presented as mean and standard error of mean, unless otherwise stated. Interrater reliability was assessed using the Cohen κ statistic. A DISCERN score of +1 or –1 point was considered agreement for each category in accordance with previous studies. Data was assessed for normality using the Shapiro-Wilk test. The one-way ANOVA and Kruskal-Wallis tests were used to assess the relationship between categorical variables, such as channel type, and the DISCERN scores with parametric and non-parametric distributions, respectively. Associations between engagement metrics and DISCERN scores were evaluated using linear regression. Statistical significance was set at p<0.05.

## 3. Results

### 3.1. Video selection and characteristics

We identified 600 videos in total based upon the three search terms. Following de-duplication, 143 videos were removed, yielding 457 videos. After initial screening of video titles and channels, 138 videos that did not meet inclusion criteria were removed. This led to 319 videos being included in the final analysis. The video review process is illustrated in Figure 1.

**Figure 1.**
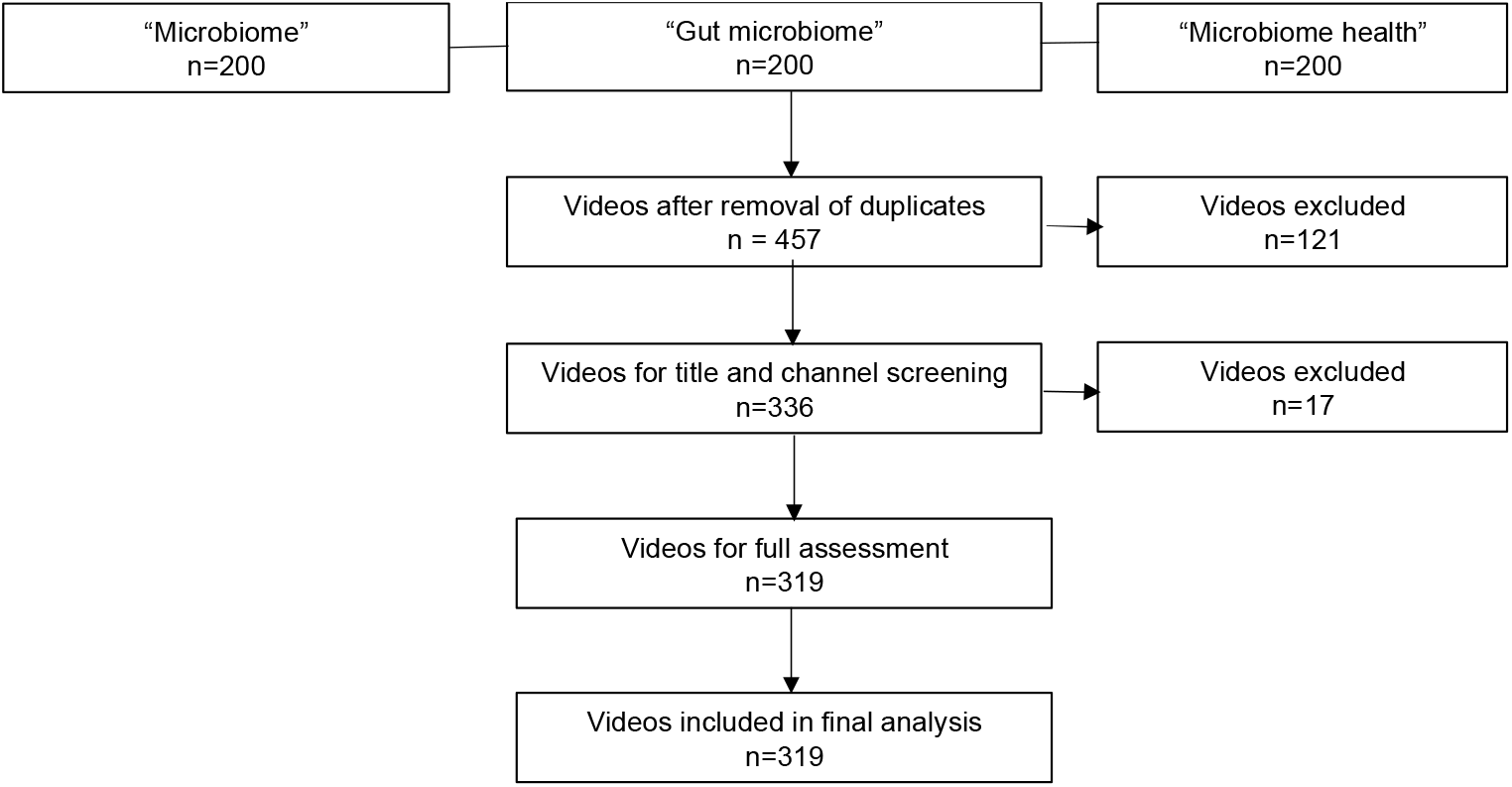
Flow diagram for the results for searches on microbiome-related videos on YouTube and the video selection process for inclusion in the study.

Of the 319 videos included in the study, the majority originated from North America (73.4%), followed by European countries (14.4%) and Australasia (10.0%). 41.0% of the videos were produced by non-profit organizations, 13.1% of videos were created by independent creators, and 11.9% were made by non-medical educational organisations. In total, only 19.4% videos were produced by a medical professional, of which a smaller minority had a proven expertise in microbiome. The mean duration of videos was 21.2 (1.25) minutes. Majority (78.6%) of videos were uploaded in the last three years, with a median age of 2.22 years. Of note, 16% of the videos had an advertisement for a product or promoted a health-related intervention for financial purposes. The characteristics of the included videos are summarized in Table 1.

**Table 1:** Characteristics of videos. All data are given as mean (standard mean) unless otherwise stated.

| Characteristic | Data point |
| --- | --- |
| Country of origin (%) |  |
| 1. Australia | 28 |
| 2. Canada | 10 |
| 3. France | 2 |
| 4. Germany | 3 |
| 5. Hungary | 1 |
| 6. India | 3 |
| 7. Ireland | 3 |
| 8. Italy | 1 |
| 9. Lebanon | 1 |
| 10. Netherlands | 1 |
| 11. New Zealand | 4 |
| 12. Russia | 1 |
| 13. South Africa | 3 |
| 14. Spain | 1 |
| 15. Switzerland | 1 |
| 16. UK | 32 |
| 17. USA | 224 |
| Channel type |  |
| 1. Educational (Non-medical) | 38 |
| 2. Educational (Medical) | 62 |
| 3. Independent Users | 49 |
| 4. Internet Media | 32 |
| 5. News Agency | 7 |
| 6. Non-Profit | 131 |
| Duration of video in minutes | 21.2 (1.25) |
| Number of days since upload | 962 (40.1) |
| Engagement |  |
| 1. Views | 195469 (43985) |
| 2. Views per day since upload | 245 (47.8) |
| 3. Likes | 3954 (909) |
| 4. Dislikes | 90.8 (21.2) |
| Number of videos with advertisement/promotional purpose | 52 |

### 3.2. Quality assessment scores

The average DISCERN and HONcode scores were 49.5 (0.68) out of 80 and 5.05 (2.52) out of 8, respectively. Of the various characteristics, regression analysis notes that only the channel type and the presence of promotional material were indicative of content quality, as per DISCERN scoring (Figure 2). Videos with promotional material scored significantly lower DISCERN scores than videos without any advertisements or product promotion (p<0.01). The highest DISCERN scores were recorded for videos by medical professionals (53.2 (0.17)) whereas the lower DISCERN scores were recorded for videos by independent content creators (39.1 (5.58)). Scores for each of the DISCERN sub-sections (completeness, understandability, relevance, depth, and accuracy of information) followed a similar trend.

**Figure 2.**
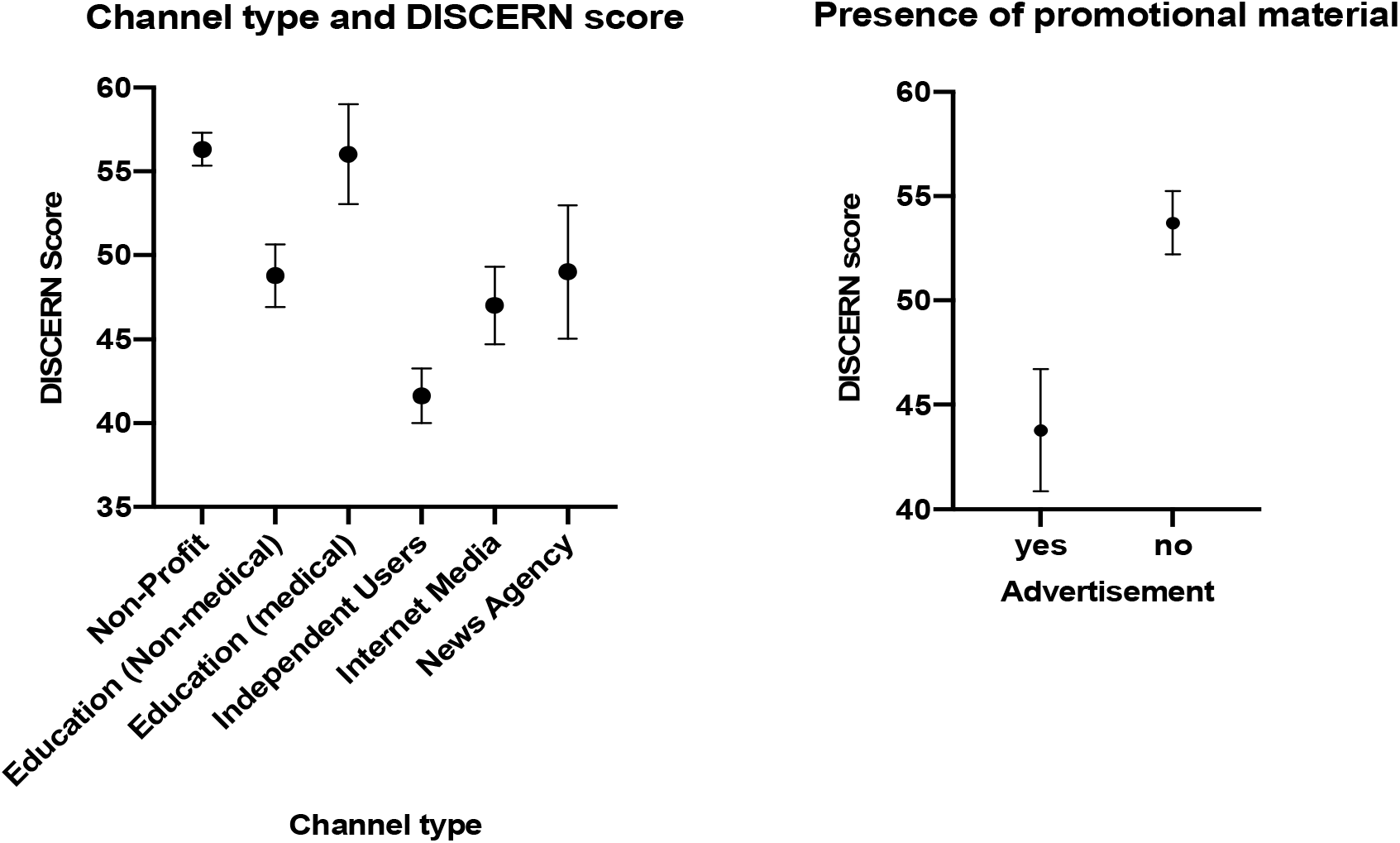
: Relationship between video characteristic and DISCERN scores. 2a (left) shows the effect of channel type on the mean DISCERN score; 2b (right) shows effect of presence of promotional material on mean DISCERN score. All values are presented as averages with 95% confidence intervals.

Linear regression analysis revealed no correlation between the quality of video based on DISCERN score and any engagement metric, including total viewership, daily viewership and preferred viewership (number of likes) (Figure 3). There was some agreement between the HONcode and DISCERN scores. For example, the HONcode score was highest for videos by medical professionals, which also had the highest DISCERN scores. However, this was not a consistent correlation as HONcode scores were significantly lowest for videos by internet media sources at 1.28 (0.81), relative to the DISCERN score of 47.7 (4.18).

**Figure 3.**
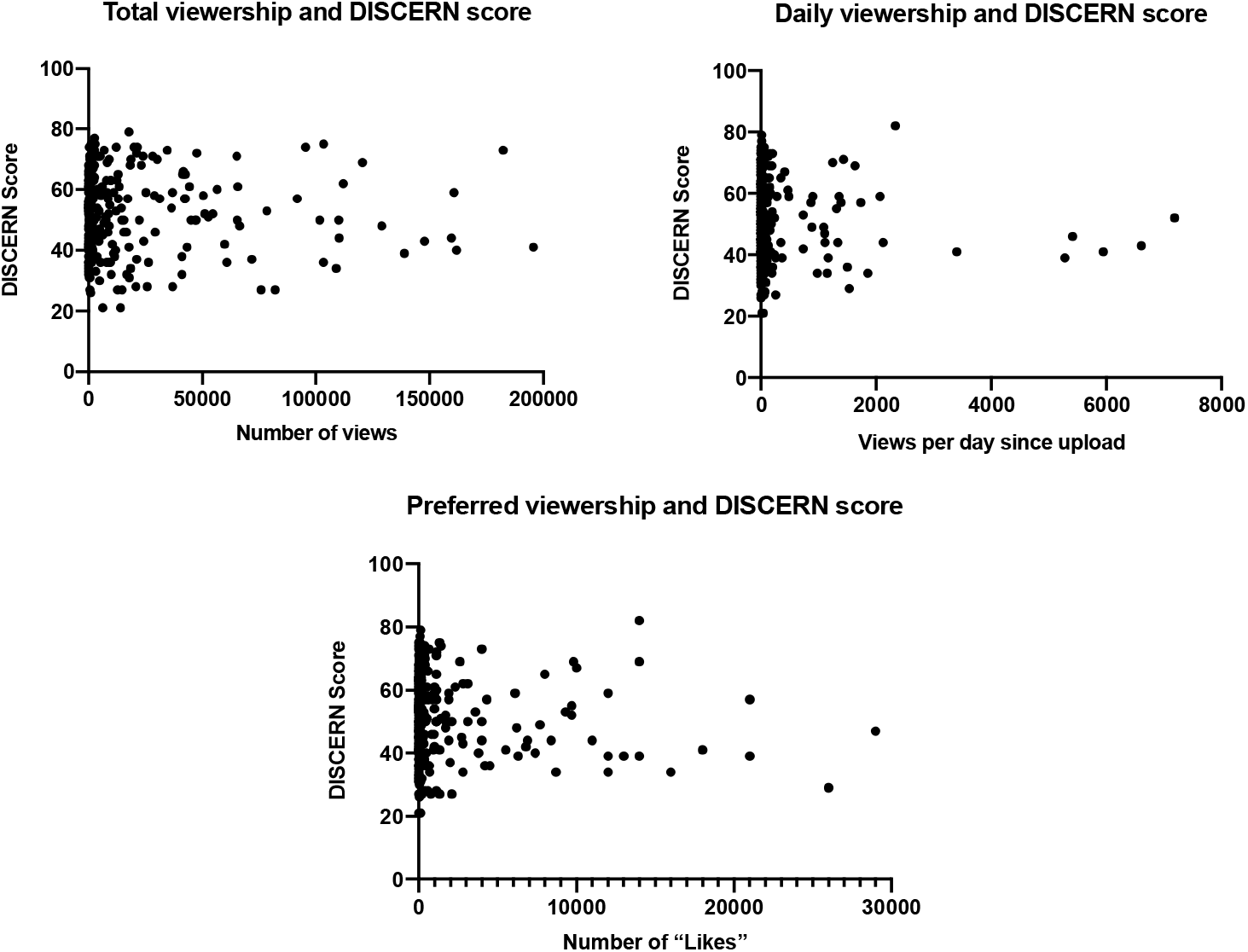
: Relationship between engagement metrics and DISCERN score, highlighting the correlation between total number of views (3a, top left), daily viewership (3b top right) and number of likes (bottom center).

### 3.3. Engagement metrics

Overall, the videos accrued a total of 62,354,628 views since their upload. The highest mean number of views was achieved by non-medical educational creators. The viewership per video for content produced by medical professionals was 136,606, which was significantly lower than the average of 195,469 views. Preferred viewership followed a similar trend, in which the highest values of 10,920 “likes” were registered for videos made by non-medical educational creators, which was three times the value for videos by healthcare professionals (n=3,480) and five times the videos from non-profit organisations (n=2,954). In contrast, the daily viewership (n=1,067) was the highest for internet media sources and was thrice that achieved by medical professionals (n=321). In parallel, videos by internet media sources were liked (n=6,020) to a greater extent relative to total viewership and compared to other channel types, reflecting a greater approval amongst the audience. Summary data of engagement metrics are included in Table 2.

**Table 2:** Engagement metrics, adherence to the Health on the Net Foundation Code of Conduct (HONCode), and DISCERN scores for videos on microbiome

| Parameters | Educational<br>(nonmedical) | Educational<br>(medical) | Independent<br>nonmedical<br>users | Internet<br>media | News<br>agencies | Non-profit<br>organizations | Overall |
| --- | --- | --- | --- | --- | --- | --- | --- |
| Videos | 38 | 62 | 49 | 32 | 7 | 131 | 319 |
| Total views | 16,786,496 | 8,469,633 | 5909797 | 3014580 | 727,969 | 27,446,153 | 62,354,628 |
| Views per<br>video | 441,749 | 136,606 | 120,608 | 94,205 | 103,995 | 209,512 | 1,106,675 |
| Daily Views | 436 | 321 | 234 | 1067 | 40 | 156 | 2,254 |
| Likes | 10920 | 3480 | 3196 | 6020 | 1162 | 2954 | 27,732 |
| Dislikes | 228 | 31 | 54 | 245 | 17 | 98 | 673 |
| HONcode<br>adherence | 2.36 (0.16) | 2.77 (0.8) | 2.48 (0.35) | 2.03<br>(0.17) | 1.28<br>(0.81) | 2.67 (0.13) | 5.05 (2.52) |
| DISCERN<br>score | 46.4 (1.85) | 53.2 (0.17) | 39.1 (5.58) | 44.9<br>(2.25) | 47.7<br>(4.18) | 53.6 (0.91) | 49.5 (0.68) |
| Reliability | 24.6 (0.96) | 27.4 (0.8) | 18.5 (2.65) | 22.6<br>(1.23) | 22.6<br>(2.29) | 27.6 (0.47) | 25.2 (0.38) |
| Treatment<br>quality | 18.9 (0.91) | 22.3 (0.63) | 18.1 (2.58) | 19.6<br>(1.02) | 21.7<br>(0.35) | 22.4 (0.45) | 21.0 (0.31) |
| Overall<br>quality | 2.84 (0.1) | 3.51 (0.09) | 2.51 (0.35) | 2.75<br>(0.16) | 3.42<br>(0.35) | 3.61 (0.06) | 3.24 (0.05) |

## 4. Discussion

This study highlights the importance of YouTube as a medium for sharing content related to the human gut microbiome. We demonstrate increasing public interest in this field, with over 62 million cumulative views on the videos that were shortlisted. However, our study also highlights that there is significant variation in the quality of information provided by most videos. Content analysis revealed that the quality of information was dependent on factors such as the profile of content creator and the presence of a financial slant. Interestingly, videos with the most credible information did not consistently receive the highest ratings or engagement. The disconnect between information quality and public engagement suggests that less informed videos are being viewed in lieu of more balanced content, which could lead to the propagation of information disorder.

Our analysis showed an inverse relationship between channel type, information quality and viewership. Videos created by recognized institutions such as universities, hospitals and charities provided more credible information than independent producers, some of whom were also qualified medical practitioners. However, both the total and average viewership was significantly higher for non-medical content creators, even after accounting for duration of videos and date of upload. The number of “likes” given to videos reflected a similar pattern, indicating that viewers expressed a more positive reception of videos that were of a lower credibility. This highlights the alarming phenomenon that low-quality, unreliable and potentially misinformed content may be more readily acquired, processed and well regarded by audience who may otherwise be unaware of existing literature on the microbiome (17).

Furthermore, the majority of videos made by independent creators captured in this study usually promoted a specific form of healthcare intervention, such as a dietary product, in order to re-engineer the microbiome (18). Although the microbiome is inarguably crucial in a wide range of common medical conditions, there is a paucity of high-grade evidence demonstrating a risk modification effect of a re-configured microbiome, or that the advertised intervention modified the microbiome to a clinically significant extent. These videos, often with no declaration of financial interests, frequently lacked evidence or misrepresented a significant leap in the interpretation of results of existing literature on the microbiome. Thus, given the far-reaching effects of online misinformation, there is a pressing need for key stakeholders, such as content creators, governments, health organisations and hosting platforms, to proactively implement policy, regulatory and educational interventions in order to protect susceptible members of the population.

### Policy Implications

Particularly in the post-pandemic era, digital technologies are commonly used in the delivery of public health and wellbeing educational material and interventions. Although younger cohorts may have embraced this shift, there is evidence to suggest that many segments of the population were poorly prepared for this rapid digitisation in services (19). Ensuring equity across the population in the ability to access, interpret and appraise information is commonly over-looked. Targeted interventions need to (1) empower all potential end-users as well as (2) serve as a custodial role over the production of content. As such, these strategies can be established at three levels in the digital information ecosystem: governmental, industry and consumer focused.

At the governmental level, there has been laudable tightening in protective legislation globally. The proposed UK legislation (the Online Safety Bill) aims to combat harmful online content through increased regulatory authority oversight (20). Similarly in the US, the proposed Health Misinformation Act and Justice Against Malicious Algorithms Act aim to hold online platforms accountable for content with misinformation related to an existing public health emergency (i.e. COVID-19) or content that contributes towards the physical or severe emotional injury of viewers (21–23).

In the context of COVID-19 and anti-vaccination misinformation, technology corporations have begun to remove and monitor non-factual and harmful content on their sites which have previously led to to passive or incidental reinforcement loops of low-quality health information. For example, YouTube’s search algorithm has historically recommended videos that attracted the most views or clicks. However, the recent heightened concerns regarding harmful misinformation on YouTube has prompted algorithm changes, which have reportedly reduced consumption of borderline content by 70% (24). Further potential action through increasing the ranking and visibility of health content from reputable scientific sources such as universities, hospitals and health charities would improve consumer exposure to high-quality and reliable health information. Platforms can also necessitate that content creators disclose potential conflicts of interest to minimise instances where content is skewed for the purpose of commercialisation. Whether it is through in-house teams or third-party organisations, social media platforms should also look to quality appraise the content they host, in addition appraising the credibility of the source. Although the temptation for big tech companies would be to employ data-driven approaches, such as through machine learning systems, it is also important to ensure an appropriate level of human oversight in this process. Similar to “likes” and “ratings”, social media platforms can employ verified users who have prior experience in using validated tools to assess video quality to provide their feedback. Videos identified can be checked by the platform before consideration for removal as well. It is important that the crude censorship of these videos does not foray into impeding the autonomy for free speech, and still retains the user at the heart of the consumption process to make the final decision.

Consumers also have a responsibility in spotting non-factual information and discerning high-quality and reliable information online. Often, social media allows users to tailor their preferences and see information from only the sources they select, leading to “bubbles” and “echo chambers” that reinforce any false information they encounter. Given the vast amount of content that is uploaded online on a continuous basis, it is unrealistic for content hosts to review all material for non-factual or harmful content. Thus, the provision of educational material for the general public is crucial, especially given that health information resources are increasingly being migrated online. Globally, the World Health Organization, in partnership with local government agencies, have introduced several initiatives to improve public awareness and education regarding online health misinformation, with a particular focus on COVID-19 and vaccination misinformation (25,26). Additionally, the UK government has recently published its Online Media Literacy Strategy, with one of the key aims being to improve the skill of the general public in identifying misinformation and assessing the reliability of an online information source (27,28). With an increasing reliance on digital health technology that is only expected to rise in the future, it is critical to ensure the public are adequately equipped with the skills to use it, and the ability to recognise and manage misinformation forms a significant component of this.

### Strengths and limitations

Our study has several strengths. Firstly, this is the first study to report on the credibility and quality of the most widely viewed YouTube videos about the gut microbiome. Furthermore, we have analysed the information quality using a combination of objective measures, including the DISCERN criteria. Our sample size is extensive and covers up to 400 most watched videos, capturing over 62 million views. As such, our study provides a broad assessment of the mismatch between information quality and viewership, which provides important insights into future discussions through which key stakeholders may co-design interventions to deliver high-quality information to an evidently receptive audience. Limitations of our study include inherent selection bias given that only videos in the English language were included, which reduces the generalisability of our results to other languages and geographical regions. In addition, our search was limited to the YouTube platform. Other social media platforms such as Facebook, TikTok, Instagram and Twitter were not included in our study, although these remain a target of high usage as well.

## 5. Conclusion

There is a significant degree of variation in the credibility of health-related YouTube videos on the gut microbiome. Both channel type and presence of financial intent were significant factors in the quality, credibility and transparency of information provided. There is little correlation between viewership and information quality, reflecting a mismatch in public engagement and discernment of good-quality health advice from misinformation. This calls for a greater scrutiny of health-related information provided on social media platforms. Further work should aim to impose more stringent regulations, policies, and educational resources to ensure accurate and reliable information is accessible in a transparent manner with the interests of the general public as the primary priority.

## Data Availability

All data produced in the present work are contained in the manuscript

## Contributors

SC, CC and VS conceived the idea for the study. SC, CC and VS designed the study. SC, YM, and VS were involved in data collection. SC, CC and YM analysed and interpreted the data. SC, VS, SRM, JA and AD drafted the manuscript. SC and VS are the study guarantors. All authors reviewed the final manuscript, agreed to be accountable for all aspects of the work, and approved the final manuscript for submission. The corresponding author attests that all listed authors meet authorship criteria and that no others meeting the criteria have been omitted.

## Ethical approval

Not required.

## Data sharing

Not applicable.

## Patient and public involvement

Patients were not invited to comment on the study design and were not consulted to develop patient relevant outcomes or interpret the results.

